# Dispensaries and medical marijuana certifications and indications: Unveiling the geographic connections in Pennsylvania, USA

**DOI:** 10.1101/2023.08.14.23294072

**Authors:** Annemarie G. Hirsch, Eric A. Wright, Cara M. Nordberg, Joseph DeWalle, Elana L. Stains, Amy L. Kennalley, Joy Zhang, Lorraine D. Tusing, Brian J. Piper

**Affiliations:** Department of Population Health Sciences, Geisinger, Danville, PA; Center for Pharmacy Innovation and Outcomes, Geisinger, Danville, PA; Geisinger Commonwealth School of Medicine, Scranton, PA; University of Pennsylvania, Philadelphia, PA

**Keywords:** medical cannabis, geospatial, evidence-based medicine, marijuana, dispensary

## Abstract

**Background:** Pennsylvania opened its first medical marijuana (MMJ) dispensary in 2018. Qualifying conditions include six conditions determined to have insufficient evidence to support or refute MMJ effectiveness. We conducted a study to describe MMJ dispensary access in Pennsylvania and to determine whether dispensary proximity was associated with MMJ certifications and community demographics.

**Methods:** Using data from the Pennsylvania Department of Health, we geocoded MMJ dispensary locations and linked them to U.S. Census Bureau data. We created dispensary access measures from the population-weighted centroid of Zip Code Tabulation Areas (ZCTAs): distance to nearest dispensary and density of dispensaries within a 15-minute drive.

We evaluated associations between dispensary access and the proportion of adults who received MMJ certification and the proportion of certifications for insufficient evidence conditions (amyotrophic lateral sclerosis, epilepsy, glaucoma, Huntington’s disease, opioid use disorder, and Parkinson’s disease) using negative binomial modeling, adjusting for community features. To evaluate associations between the proportion of the population that was non-White, Hispanic, or both (NW-H) and distance to nearest dispensary, we used logistic regression to estimate the odds ratios (OR) and 95% confidence intervals (CI), adjusting for median income.

**Results:** Distance and density of MMJ dispensaries was associated with the proportion of the ZCTA population certified and the proportion of certifications for limited evidence conditions. Compared to ZCTAs with no dispensary within 15 minutes, the proportion of adults certified increased by up to 31% and the proportion of certifications for limited evidence decreased by up to 22% for ZCTAs with two dispensaries. In 2021, the odds of being within five miles of a dispensary was higher in ZCTAs with the highest proportions of NW-H individuals (OR: 26.05, CI: 16.7 - 40.6), compared to ZCTAs with the lowest proportions.

**Conclusions:** Greater dispensary access was associated with the proportions of certified residents and certifications for insufficient evidence conditions. Whether these patterns are due to differences in accessibility or demand is unknown. Associations between community demographics and dispensary proximity may indicate MMJ access differences.

## Introduction

The legalization of medical marijuana (MMJ) is expanding worldwide, including in some parts of the U.S., Europe, Asia, and Africa.^1^ In the U.S., as of 2023, 38 states allow medical use of cannabis products.^2^ States have a growing list of qualifying conditions for MMJ, despite limited evidence of the effectiveness of MMJ for many of these conditions.^3^ The geographic location of MMJ dispensaries has been associated with marijuana use,^4–6^ however it is unknown whether the locations of MMJ dispensaries is associated with the qualifying conditions for which individuals are being certified. As MMJ legalization and the number of certifying conditions in the U.S. expands,^2,7^ it is imperative to understand the potential implications of the locations of MMJ dispensaries.

Geographic locations of MMJ dispensaries have been associated with marijuana use patterns. Living near a higher number of MMJ dispensaries has been associated with a greater number of days of marijuana use, greater marijuana demand, and frequency of marijuana use.^4–6^ Much of this research has been conducted in California, the first state to legalize MMJ in 1996. It is unknown whether these findings are generalizable to states in other regions of the country.

Very little is known about whether geographic access to MMJ dispensaries is associated with the types of qualifying conditions for which people are certified. In 2017, the National Academies of Sciences, Engineering and Medicine (NASEM) published a comprehensive review of the evidence regarding the health effects of using cannabis and cannabis-derived products.^3^ In the report, NASEM categorized conditions into one of five categories: conclusive evidence, substantial evidence, moderate evidence, limited evidence, and no or insufficient evidence to support the association.^3^ A subsequent report identified a mismatch between many of the qualifying conditions allowed under state law and the evidence supporting the use of MMJ. In 2019, a national report estimated 15.4% of patient-reported qualifying conditions had less than substantial evidence of the effectiveness of MMJ treatment.^8^ Since that time, new states and new qualifying conditions have been added to MMJ regulations.^7^

Different community characteristics have been associated with geographic access to MMJ dispensaries, but results have differed across states. In New York state, for example. MMJ services were least available in neighborhoods with highly educated residents,^9^ while in Oklahoma census tracts with at least one MMJ dispensary had a higher proportion of uninsured individuals living below the poverty level.^10^ In New York state, MMJ services were least available in neighborhoods with Black residents,^9^ while in Los Angeles the presence of MMJ dispensaries was associated with a higher proportion of Hispanic residents.^11^ Differences in geographic proximity to MMJ dispensaries may impact access to effective treatment options for conditions such as chronic pain or multiple sclerosis.^3^ Conversely, there is some prior evidence that closer proximity may have negative consequences, as proximity has been associated with elevated rates of marijuana-related hospitalizations and crime.^12–14^

As the number of MMJ dispensaries grow, it is important to understand the implications of where states locate MMJ dispensaries and how to provide equitable access. Pennsylvania legalized MMJ in 2016 and opened its first dispensary in 2018. Using data from the Pennsylvania Department of Health, we conducted a study of the association between proximity to MMJ dispensaries and both the proportion of individuals certified and the proportion of certifications for conditions that have no or insufficient evidence. We then evaluated the association between racial, ethnic, socioeconomic community features and access to Pennsylvania dispensaries.

## Methods

We conducted a cross-sectional study of MMJ dispensaries in Pennsylvania zip code tabulation areas (ZCTAs) from 2018 to 2021 using data from the Pennsylvania Department of Health obtained 2022 and the Census Bureau and American Community Survey. According to the Census Bureau, Pennsylvania was the fifth most populous state in 2020 in the US (population = 13.0 million, 26.6% non-White). We evaluated associations between geographic access, defined using distance and density measures, to MMJ dispensaries and certifications. We then measured associations between community sociodemographic factors and MMJ dispensary access.

### Measures of Geographic Access

The Pennsylvania Department of Health provided the locations and opening dates of MMJ dispensaries in Pennsylvania. Using ArcGIS V.10.8 (ESRI, Redlands, California) we geocoded dispensary locations for each year between 2018 and 2021 and created two dispensary access measures from the population-weighted centroid of ZCTAs with at least 100 adult residents: Geodesic distance measure (miles) to nearest dispensary and a density measure of the count of dispensaries within a 15-minute driving radius (0, 1, 2) using ArcGIS Network Analyst and StreetMap Premium 2021 release 1 dataset.

### Measures of MMJ use

For each year between 2018 and 2021, the Pennsylvania Department of Health provided certification data at the Zip Code level to Pennsylvania Academic Clinical Research Centers in February 2023. Data included 5-digit zip code for the certifying person, certification status (included: active, inactive, pending, expired, cancelled), creation date of certification, treatment period (by number of months up to 12) and up to 10 qualifying serious medical conditions approved by the Department of Health. Zip Code to ZCTA crosswalk files from UDS Mapper (HRSA, 2018 – 2021) were used to summarize MMJ certifications at the ZCTA level. For each ZCTA, we calculated the proportion of adults residing in a ZCTA who had a certification in each year and the proportion of certifications for insufficient evidence conditions, per the NASEM report. To calculate the proportion certified, we divided the number of certifications in each year between 2018 and 2021 by the size of the adult population in that ZCTA using data from the U.S. Census Bureau and the American Community Survey data.

We identified six conditions on the Pennsylvania’s list of qualifying conditions between 2019 – 2021 that the NASEM categorized as having no or insufficient evidence of MMJ effectiveness^2^ (insufficient evidence conditions): myotrophic lateral sclerosis, epilepsy, glaucoma, Huntington’s disease, opioid use disorder, and Parkinson’s disease (**Supplement Table 1**). Because Pennsylvania expanded their list of qualifying conditions in 2019, we calculated the proportion of certifications that were only for one or more of these conditions for each year between 2019 and 2021 by dividing the number of certifications for the six insufficient evidence conditions by the total number of certifications.

### Community Measures

Using data from the American Community Survey, we created measures of income and racial and ethnic composition for each of the ZCTAs. For each year, we quartiled the median household income and the proportion of residents who were non-White, Hispanic, or both (NH-W). We categorized ZCTA’s by level of urbanicity using the Rural-Urban Commuting Area Codes from the US Department of Agriculture based on Census Bureau data.^15^ ZCTA’s were categorized as metropolitan, micropolitan/small town, and rural.

### Analysis

The goals of these analyses were to describe MMJ location and certification patterns in Pennsylvania; evaluate associations between geographic access to MMJ dispensaries (distance and density) and two MMJ use outcomes: proportion of population certified and proportion of certifications for insufficient evidence conditions; and evaluate associations between community features and MMJ access. We evaluated associations between dispensary access (distance: less than five miles (8.1 kilometers), five to fewer than ten miles (16.1 kilometers), ten and more miles; distance: 0, 1, 2 or more within a 15 minute driving radius), the proportion of adults who received MMJ certification (2018 – 2021), and the proportion of certifications for limited evidence conditions (2019 – 2021), using negative binomial modeling to estimate the incidence rate ratio (IRR) and 95% confidence intervals (CI), separately, for each outcome in each year. For these models, we used count variables (i.e., number of certifications) as the outcomes and log-transformed denominators (i.e. population and count of certifications, respectively) as offset terms. We used an unadjusted model (model 1) and then we added two sociodemographic factors, one at a time, to that model, to avoid violations of non-positivity: proportion NW-H (model 2); median income (model 3).^16^ To evaluate associations between proportion of the population that was NH-W and distance to nearest dispensary (distance: <5 miles, 5 miles or more), we used logistic regression to estimate the odds ratios (OR) and 95% CIs, adjusting for median income. Urbanicity was not included in final models given that, as of 2021, no ZCTAs in Pennsylvania categorized as rural had a MMJ dispensary.

## Results

Of 1,831 ZCTAs, 1,608 were included in the analysis. From 2018 to 2021, the median distance to the nearest dispensary decreased from 14.7 miles to 9.3 miles (23.7 kilometers to 15.0 kilometers). The percent of ZCTAs within five miles of a dispensary nearly doubled, from 16.5% to 30.5%. The percent of ZCTAs with at least two dispensaries within 15 minutes more than tripled, from 9.2% to 27.8% (**Figure 1**, **Table 1**). The median percent of population certified for MMJ in Pennsylvania ZCTAs increased from 0.55% to 3.54% (**Figure 1**) and the median percent of certifications for low evidence conditions decreased from 3.3% to 1.9%.

**Figure 1.**
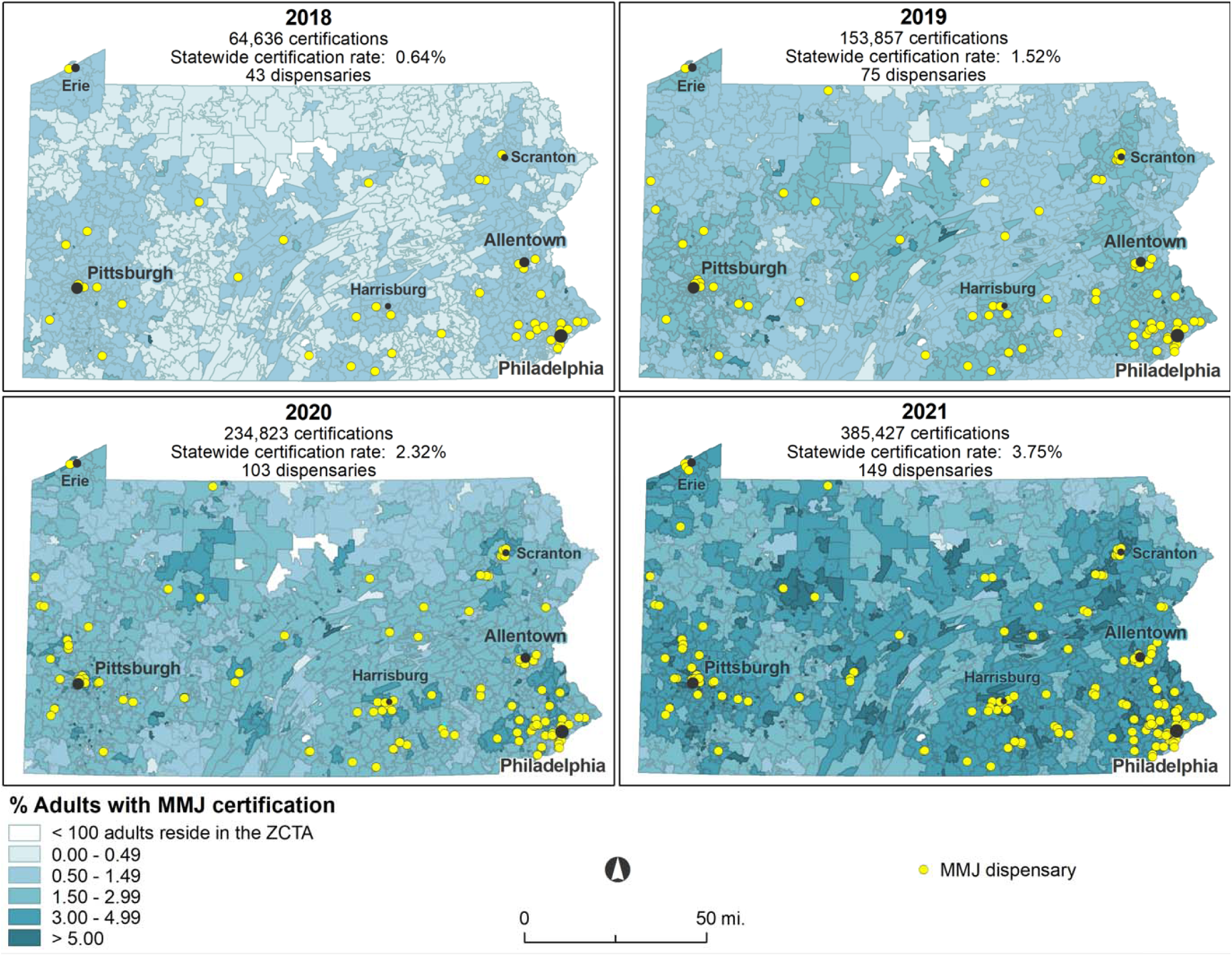
Pennsylvania maps of percent of adults with medical marijuana (MMJ) certification 2018 – 2021 in Zip Code Tabulation Areas with at least 100 adult residents.

**Table 1.** Descriptions of distance and density to medical marijuana dispensaries and medical marijuana certifications in Pennsylvania Zip Code Tabulation Areas^1^ 2018 – 2021.

|  | 2018 |  |  | 2019 |  |  | 2020 |  |  | 2021 |  |  |
| --- | --- | --- | --- | --- | --- | --- | --- | --- | --- | --- | --- | --- |
| **Distance to nearest dispensary (miles)** | ≤ 5 | >5 to ≤10 | >10 | ≤ 5 | >5 to ≤10 | >10 | ≤ 5 | >5 to ≤10 | >10 | ≤ 5 | >5 to ≤10 | >10 |
| n (%) | 266 (16) | 287 (18) | 1055 (66) | 356 (22) | 323 (20) | 928 (58) | 418 (26) | 371 (23) | 819 (51) | 491 (31) | 357 (22) | 760 (47) |
| Percent certified: mean (sd) | 0.8 (0.5) | 0.7 (0.5) | 0.5 (0.3) | 1.7 (0.7) | 1.7 (0.8) | 1.3 (0.8) | 2.6 (1.0) | 2.3 (1.0) | 1.9 (1.1) | 4.2 (1.5) | 3.8 (1.6) | 3.1 (16.4) |
| Percent certified for insufficient evidence conditions<sup>2</sup>: mean (sd) |  |  |  | 4.1 (3.5) | 4.3 (4.8) | 6.2 (10.9) | 2.9 (2.6) | 3.2 (4.3) | 4.2 (6.4) | 2.2 (2.2) | 2.7 (4.4) | 3.3 (4.4) |
| **Density within 15 minute drive** | 0 | 1 | ≥2 | 0 | 1 | ≥2 | 0 | 1 | ≥2 | 0 | 1 | ≥2 |
| n (%) | 1230 (76) | 230 (14) | 148 (9) | 1091 (68) | 268 (17) | 249 (15) | 1007(63) | 263 (16) | 338 (21) | 926 (58) | 235 (15) | 447 (28) |
| Percent certified: mean (sd) | 0.5 (0.4) | 0.8 (0.6) | 0.8 (0.5) | 1.3 (0.8) | 1.7 (0.9) | 1.7 (0.8) | 1.9 (1.1) | 2.5 (1.1) | 2.6 (1.0) | 3.2 (1.6) | 4.1 (1.6) | 4.1 (1.6) |
| Percent certified for insufficient evidence conditions<sup>2</sup>: mean (sd) |  |  |  | 6.0 (10.2) | 4.5 (4.4) | 3.7 (2.8) | 4.0 (6.1) | 3.2 (4.0) | 2.8 (2.4) | 3.2 (4.5) | 2.6 (3.5) | 2.2 (2.4) |
| **Distance to nearest MMJ dispensary (miles)** |  |  |  |  |  |  |  |  |  |  |  |  |
|  | 2018 |  |  | 2019 |  |  | 2020 |  |  | 2021 |  |  |
| Percent non-White, Hispanic, or both | Mean (sd) |  |  | Mean (sd) |  |  | Mean (sd) |  |  | Mean (sd) |  |  |
| Quartile 1 (lowest %) | 22.8 (11.7) |  |  | 17.8 (8.8) |  |  | 16.8 (9.0) |  |  | 15.9 (8.6) |  |  |
| Quartile 2 | 21.0 (12.4) |  |  | 17.2 (10.5) |  |  | 16.5 (10.7) |  |  | 15.4 (10.7) |  |  |
| Quartile 3 | 15.7 (11.3) |  |  | 12.6 (10.0) |  |  | 11.3 (9.8) |  |  | 10.5 (9.6) |  |  |
| Quartile 4 (highest %) | 10.0 (10.6) |  |  | 8.0 (8.8) |  |  | 5.9 (6.6) |  |  | 5.4 (6.6) |  |  |
| Median income |  |  |  |  |  |  |  |  |  |  |  |  |
| Quartile 1 (lowest %) | 19.9 (13.9) |  |  | 15.4 (11.4) |  |  | 14.1 (11.4) |  |  | 13.3 (11.3) |  |  |
| Quartile 2 | 21.7 (12.9) |  |  | 17.0 (10.7) |  |  | 16.1 (10.8) |  |  | 15.0 (10.6) |  |  |
| Quartile 3 | 17.0 (10.9) |  |  | 14.2 (9.7) |  |  | 12.8 (9.4) |  |  | 12.0 (9.2) |  |  |
| Quartile 4 (highest %) | 10.8 (9.1) |  |  | 9.0 (7.4) |  |  | 7.4 (6.2) |  |  | 6.7 (6.1) |  |  |

<sup>1</sup>1608 Zip Code Tabulation Areas with at least 100 adult residents in every year from 2018 – 2021. <sup>2</sup>Qualifying conditions that the National Academies of Sciences, Engineering, and Medicine indicates there is no or insufficient evidence to support or refute treatment effectiveness of medical marijuana: amyotrophic lateral sclerosis, epilepsy, glaucoma, Huntington's disease, opioid use disorder, Parkinson's disease. Abbreviations: sd: standard deviation

In unadjusted and adjusted models, the proportion of the population certified increased with greater dispensary access (i.e., shorter distance, higher density) (**Table 2**). This finding was present in each year from 2018 to 2021. Compared to ZCTAs with the nearest dispensary more than ten miles away, the proportion of adults certified increased by up to 47% for ZCTAs within 5 to 10 miles of a dispensary and 42% for ZCTAs within 5 miles (2018, model 3). Compared to ZCTAs with zero dispensaries within a 15 minute drive, the proportion of adults certified increased by up to 35% for ZCTAs with at least 1 dispensary and 31% for ZCTAs with at least 2 dispensaries (2018, model 1). These associations remained after adjusting for community features (models 2 and 3).

**Table 2.** Associations between geographic access to medical marijuana dispensaries in Zip Code Tabulation Areas (ZCTA)^1^ and proportion of certified adults in Pennsylvania^2^.

**Model 1: Unadjusted associations**
|  | 2018 | 2019 | 2020 | 2021 |
| --- | --- | --- | --- | --- |
| Incident Rate Ratio (95% confidence interval) |  |  |  |  |
| **Model 1; Unadjusted associations. Distance and density modeled separately<sup>2</sup>** |  |  |  |  |
| **Distance to nearest dispensary (miles)** |  |  |  |  |
| ≤5 | 1.45 (1.37, 1.55) | 1.07 (0.99, 1.16) | 1.13 (1.05, 1.22) | 1.09 (1.02, 1.17) |
| >5 to ≤10 | 1.43 (1.36, 1.52) | 1.23 (1.16, 1.30) | 1.21 (1.16, 1.27) | 1.17 (1.12, 1.23) |
| >10 (reference) |  |  |  |  |
| **Density w/in 15 minutes** |  |  |  |  |
| 0 (reference) |  |  |  |  |
| 1 | 1.35 (1.27, 1.44) | 1.15 (1.08, 1.22) | 1.16 (1.09, 1.24) | 1.13 (1.06, 1.21) |
| ≥2 | 1.31 (1.22, 1.41) | 0.96 (0.88, 1.05) | 1.06 (0.98, 1.14) | 1.04 (0.97, 1.11) |
| **Model 2: Adjusted for median income (quartiled): Distance and density modeled separately<sup>2</sup>** |  |  |  |  |
| **Distance to nearest dispensary** |  |  |  |  |
| ≤5 | 1.39 (1.30, 1.48) | 1.13 (1.07, 1.20) | 1.15 (1.08, 1.23) | 1.10 (1.03, 1.18) |
| >5 to ≤10 | 1.32 (1.25, 1.41) | 1.16 (1.10, 1.22) | 1.13 (1.07, 1.20) | 1.12 (1.06, 1.18) |
| >10 (reference) |  |  |  |  |
| **Density w/in 15 minutes** |  |  |  |  |
| 0 (reference) |  |  |  |  |
| 1 | 1.29 (1.21, 1.37) | 1.14 (1.08, 1.20) | 1.16 (1.09, 1.23) | 1.11 (1.04, 1.19) |
| ≥2 | 1.24 (1.16, 1.33) | 1.04 (0.98, 1.10) | 1.09 (1.03, 1.15) | 1.05 (0.99, 1.12) |
| **Model 3: Adjusted for proportion non-White or Hispanic (quartiled): Distance and density modeled separately<sup>2</sup>** |  |  |  |  |
| **Distance to nearest dispensary** |  |  |  |  |
| ≤5 | 1.47 (1.37, 1.57) | 1.21 (1.13, 1.29) | 1.28 (1.21, 1.35) | 1.24 (1.18, 1.30) |
| >5 to ≤10 | 1.42 (1.33, 1.50) | 1.28 (1.21, 1.35) | 1.26 (1.20, 1.31) | 1.20 (1.15, 1.26) |
| >10 (reference) |  |  |  |  |
| **Density w/in 15 minutes** |  |  |  |  |
| 0 (reference) |  |  |  |  |
| 1 | 1.32 (1.23, 1.41) | 1.20 (1.13, 1.27) | 1.23 (1.16, 1.30) | 1.18 (1.12, 1.25) |
| ≥2 | 1.30 (1.20, 1.42) | 1.07 (0.98, 1.16) | 1.17 (1.10, 1.24) | 1.16 (1.11, 1.23) |

<sup>1</sup>1608 ZCTAs with at least 100 adult residents in every year from 2018 – 2021. <sup>2</sup>Negative binomial models separate for each outcome and year that used count variables (i.e., number of certifications) as the outcomes and log-transformed denominators (i.e. population) as offset terms.

In years 2019 to 2021, the proportion of certifications for no or insufficient evidence conditions decreased with greater dispensary access (**Table 4**). Compared to ZCTAs with the nearest dispensary more than ten miles away, the proportion of adults certified decreased by up to 30% for ZCTAs within 5 to 10 miles of a dispensary and 38% for ZCTAs within 5 miles (2021, model 1). Compared to ZCTAs with zero dispensaries within a 15 minute drive, the proportion of adults certified decreased by up to 34% for ZCTAs with at least 1 dispensary and 22% for ZCTAs with at least 2 dispensaries (2021, model 1). These associations remained after adjusting for community features (models 2 and 3).

**Table 3.**
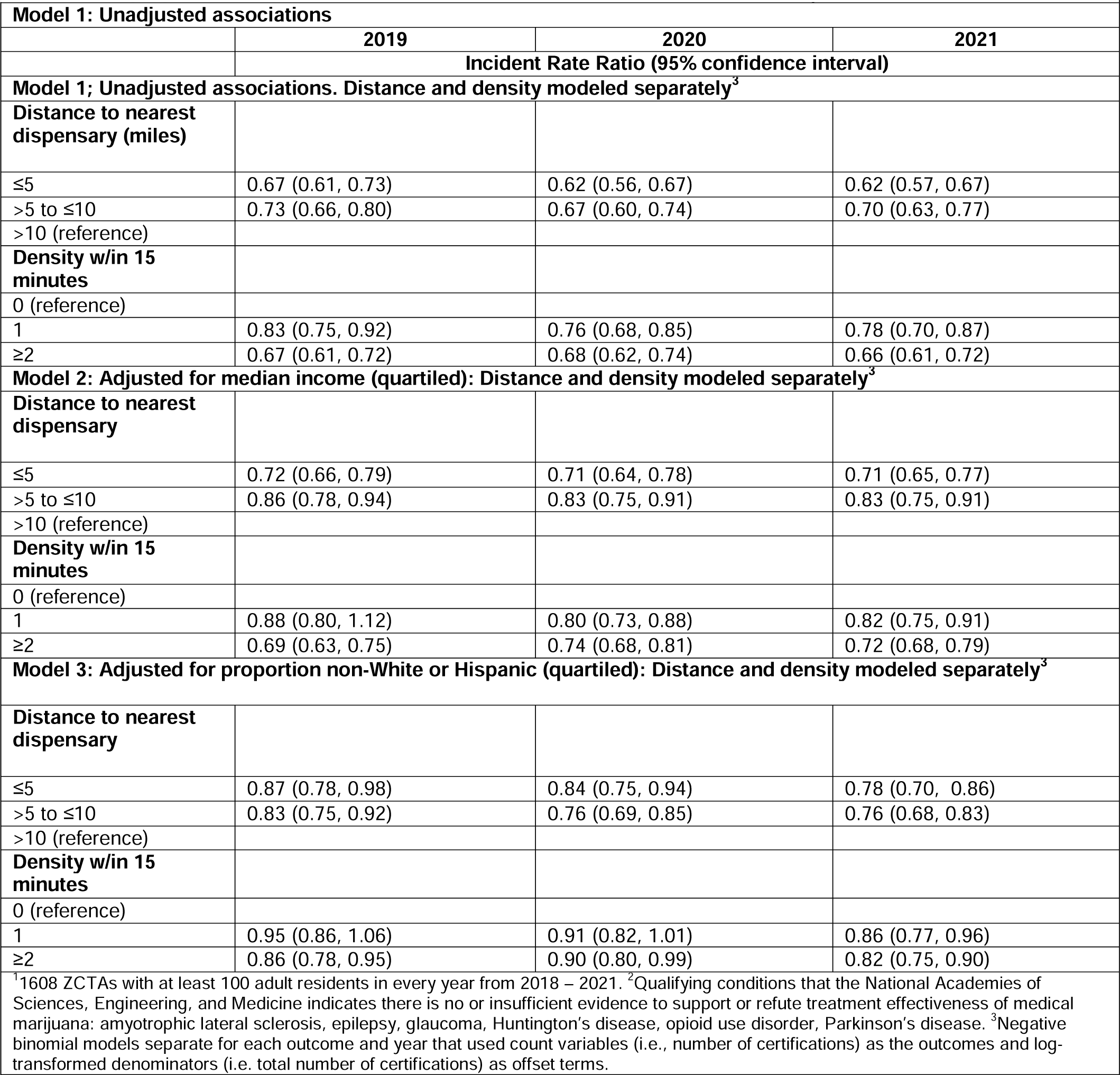
Associations between geographic access to medical marijuana dispensaries in Zip Code Tabulation Areas (ZCTA)^1^ and proportion of certifications for insufficient evidence conditions in Pennsylvania^2^.

**Model 1: Unadjusted associations**
|  | 2019 | 2020 | 2021 |
| --- | --- | --- | --- |
| **Incident Rate Ratio (95% confidence interval)** |  |  |  |
| **Model 1; Unadjusted associations. Distance and density modeled separately<sup>3</sup>** |  |  |  |
| **Distance to nearest dispensary (miles)** |  |  |  |
| ≤5 | 0.67 (0.61, 0.73) | 0.62 (0.56, 0.67) | 0.62 (0.57, 0.67) |
| >5 to ≤10 | 0.73 (0.66, 0.80) | 0.67 (0.60, 0.74) | 0.70 (0.63, 0.77) |
| >10 (reference) |  |  |  |
| **Density w/in 15 minutes** |  |  |  |
| 0 (reference) |  |  |  |
| 1 | 0.83 (0.75, 0.92) | 0.76 (0.68, 0.85) | 0.78 (0.70, 0.87) |
| ≥2 | 0.67 (0.61, 0.72) | 0.68 (0.62, 0.74) | 0.66 (0.61, 0.72) |
| **Model 2: Adjusted for median income (quartiled): Distance and density modeled separately<sup>3</sup>** |  |  |  |
| **Distance to nearest dispensary** |  |  |  |
| ≤5 | 0.72 (0.66, 0.79) | 0.71 (0.64, 0.78) | 0.71 (0.65, 0.77) |
| >5 to ≤10 | 0.86 (0.78, 0.94) | 0.83 (0.75, 0.91) | 0.83 (0.75, 0.91) |
| >10 (reference) |  |  |  |
| **Density w/in 15 minutes** |  |  |  |
| 0 (reference) |  |  |  |
| 1 | 0.88 (0.80, 1.12) | 0.80 (0.73, 0.88) | 0.82 (0.75, 0.91) |
| ≥2 | 0.69 (0.63, 0.75) | 0.74 (0.68, 0.81) | 0.72 (0.68, 0.79) |
| **Model 3: Adjusted for proportion non-White or Hispanic (quartiled): Distance and density modeled separately<sup>3</sup>** |  |  |  |
| **Distance to nearest dispensary** |  |  |  |
| ≤5 | 0.87 (0.78, 0.98) | 0.84 (0.75, 0.94) | 0.78 (0.70, 0.86) |
| >5 to ≤10 | 0.83 (0.75, 0.92) | 0.76 (0.69, 0.85) | 0.76 (0.68, 0.83) |
| >10 (reference) |  |  |  |
| **Density w/in 15 minutes** |  |  |  |
| 0 (reference) |  |  |  |
| 1 | 0.95 (0.86, 1.06) | 0.91 (0.82, 1.01) | 0.86 (0.77, 0.96) |
| ≥2 | 0.86 (0.78, 0.95) | 0.90 (0.80, 0.99) | 0.82 (0.75, 0.90) |

<sup>1</sup>1608 ZCTAs with at least 100 adult residents in every year from 2018 – 2021. <sup>2</sup>Qualifying conditions that the National Academies of Sciences, Engineering, and Medicine indicates there is no or insufficient evidence to support or refute treatment effectiveness of medical marijuana: amyotrophic lateral sclerosis, epilepsy, glaucoma, Huntington's disease, opioid use disorder, Parkinson's disease. <sup>3</sup>Negative binomial models separate for each outcome and year that used count variables (i.e., number of certifications) as the outcomes and log-transformed denominators (i.e. total number of certifications) as offset terms.

**Table 4.** Adjusted associations between community features and distance to nearest medical marijuana dispensaries in Zip Code Tabulation Areas (ZCTA)^1^.

|  | Proportion of adult ZCTA population certified |  |  |  |
| --- | --- | --- | --- | --- |
|  | 2018 | 2019 | 2020 | 2021 |
|  | Odds Ratio (95% confidence interval) |  |  |  |
| **Proportion non-White, Hispanic, or both** |  |  |  |  |
| quartile 1 (lowest) (ref) |  |  |  |  |
| quartile 2 | 1.36 (0.61, 3.03) | 2.00 (1.02, 3.90) | 1.63 (0.93, 2.84) | 2.02 (1.24, 3.28) |
| quartile 3 | 5.33 (2.71, 10.46) | 7.66 (4.23, 13.88) | 5.77 (3.52, 9.46) | 5.96 (3.81, 0.33) |
| quartile 4 (highest) | 29.61 (15.43, 56.41) | 34.93 (19.58, 62.31) | 27.30 (16.85, 44.22) | 26.05 (16.70, 40.64) |
| **Median household income** |  |  |  |  |
| quartile 1 (lowest) (ref) |  |  |  |  |
| quartile 2 | 0.67 (0.43, 1.05) | 0.71 (0.47, 1.07) | 0.68 (0.46, 1.002) | 0.64 (0.44, 0.94) |
| quartile 3 | 0.59 (0.38, 0.93) | 0.68 (0.45, 1.03) | 0.78 (0.53, 1.16) | 0.77 (0.53, 1.11) |
| quartile 4 (highest) | 0.95 (0.65, 1.40) | 0.98 (0.68, 1.42) | 1.24 (0.86, 1.77) | 1.48 (1.04, 2.10) |

<sup>1</sup>1608 ZCTAs with at least 100 adult residents in every year from 2018 – 2021.

ZCTAs with the higher proportions of NW-H individuals had higher odds of having a dispensary within five miles (versus greater than five miles) in all years than ZCTAs with the lowest proportion of NW-H individuals. Adjusting for median income, in communities with the highest proportion of NW-H (quartile 4) the odds of having a dispensary within five miles were more than 25 times the odds among communities with the lowest proportion of NW-H individuals (quartile 1) in every year. ZCTAs with higher median incomes had lower odds of having a dispensary within five miles, but most of the confidence intervals included the null value.

## Discussion

As legalization of MMJ expands worldwide,^1^ understanding the implications of the availability of MMJ in communities is essential. Geographic access to MMJ dispensaries dramatically increased in Pennsylvania from 2018 to 2021. We conducted the first study of the association between MMJ dispensary locations in Pennsylvania and MMJ certifications and the first study in the U.S. of the association between dispensary locations and qualifying conditions. We found that geographic access to MMJ dispensaries since the first dispensary opened in 2018 has consistently differed by the race and ethnic composition of Pennsylvania communities.

As of April, 2023, 38 states and Washington DC have legalized MMJ and within those states,^17^ certifications have been rapidly growing.^18^ In Pennsylvania, the proportion of adults certified for MMJ increased more than six-fold from 2018 to 2021. We observed that greater access, measured by both distance and density, was associated with MMJ certifications, independent of demographic and socioeconomic composition of the population, factors that have been associated with MMJ use.^19^ These findings are consistent with other states with a longer history of MMJ legalization. Multiple studies in California, for example, support the association between medical marijuana proximity and demand and utilization.^4–6^

In addition to the growth in the population certified, Pennsylvania has increased the number of qualifying conditions since legalization in 2016.^20^ Greater access to MMJ dispensaries was associated with the qualifying conditions for which individuals were certified. Specifically, greater distances and lower density of MMJ dispensaries were associated with a higher proportion of certifications for qualifying conditions with insufficient evidence^3^ of the effectiveness of MMJ treatment. Prior studies have demonstrated that access to care for some of these qualifying conditions, including opioid use disorder^21–22^ and epilepsy^23^, is more limited in minority racial and ethnic groups and in low income populations.^24^ However, even after adjusting for these factors, the association between distance, density, and certifications for insufficient evidence conditions remained. It may be that those communities with less access to MMJ dispensaries also have less access to specialty care and treatment that was not captured in our analyses. Limited access to traditional health care for these conditions could motivate people living in such communities to seek MMJ as an alternative treatment option.

In Pennsylvania, we observed that ZCTAs with higher proportions of NW-H residents were more likely to have a MMJ dispensary within five miles. This association was independent of median income. Studies in other states in the U.S., such as California and Colorado, have also shown the density of dispensaries to be positively associated with higher proportions of Hispanic residents^11^ and higher proportions of racial and ethnic minorities in general.^5^ Studies in Oklahoma, California and Colorado show that dispensaries are closer in proximity to communities with higher rates of poverty,^4^ higher rates of uninsured individuals,^10^ and lower income levels.^6^ Pennsylvania zoning laws (2016 Act 16) for MMJ dispensaries specify that a dispensary may not operate on the same site as a facility used for growing and processing marijuana and may not be located within 1,000 feet of the property line of a public, private, or parochial school or a day care center.^25^ Municipalities within the state have adopted a variety of zoning ordinances regarding where MMJ dispensaries can be located.^26^ The potential benefits and harms of proximity to MMJ dispensaries is still poorly understood. For individuals with conditions for which there is evidence of the effectiveness of MMJ, such as chronic pain and chemotherapy induced nausea, proximity may improve access to effective treatment options.^3^ However, some studies have reported that closer proximity is also associated with an increase in the number of marijuana hospitalizations,^13^ crime,^12^ and rates of physical abuse.^14^ Importantly, these studies demonstrate correlations, not necessarily causation. However, future research should explore the impact of ordinances on geographic access to MMJ across population subgroups, and the potential benefits and harms^27^ of proximity.

This study had some limitations. First, this was an ecological study and is vulnerable to ecological fallacy. Thus, the findings should not be interpreted as individual-level risk factors for certifications. Second, this is a cross-sectional study and it is unknown whether the association between location and certifications is due to greater geographic access or the placement of dispensaries in response to demand. There are multiple strengths to this novel study. We used two measures of geographic access to MMJ dispensaries, distance and density. In analyzing associations between geographic access and certifications we adjusted for potential community-level confounders. We evaluated these associations using data from the first four years of MMJ in Pennsylvania, a period of rapid acceleration in MMJ dispensary growth and certifications.

## Conclusions

Our study found differences in geographic access to dispensaries by the racial and ethnic composition of communities. There may be implications to where MMJ dispensaries are located, including the proportion of individuals certified for MMJ and the qualifying conditions for which they are certified. As U.S. states and countries around the world continue to consider and respond to the legalization of MMJ, it is critical to evaluate the impact of MMJ locations in the use of and access to MMJ.

## Data Availability

All data produced in the present study are available upon reasonable request to the authors

## Acknowledgements

We would like to express our gratitude to Latisha “Lolly” Bentch for making this certification data used in this study available.

## References

1. Ransing R, de la Rosa PA, Pereira-Sanchez V, et al. Current state of cannabis use, policies, and research across sixteen countries: cross-country comparisons and international perspectives. Trends in Psychiatry and Psychotherapy. 2022; 44(Suppl 1): e20210263. http://dx.doi.org/10.47626/2237-6089-2021-0263.

2. National Conference of State Legislatures. Medical Uses of Cannabis. Jun 2023. https://www.ncsl.org/health/state-medical-cannabis-laws#:∼:text=Medical%2DUse%20Update,1%20below%20for%20additional%20information. Accessed August 2023.

3. National Academies of Sciences, Engineering, and Medicine. The Health Effects of Cannabis and Cannabinoids: The Current State of Evidence and Recommendations for Research. Washington, DC: The National Academies Press; 2017. https://doi.org/10.17226/24625

4. Morrison C, Gruenewald PJ, Freisthler B, Ponicki WR, Remer LG. The economic geography of medical marijuana dispensaries in California. International Journal of Drug Policy. 2014; 25(30): 508 – 515.

5. Freisthler B, Gruenewald PJ. Examining the relationship between the physical availability of medical marijuana and marijuana use across fifty California cities. Drug and Alcohol Dependence. 2014; 143; 244 – 250.

6. Shih RA, Tucker JS, Pedersen ER, Seelam R, Dunbar MS, Kofner A, Firth C, D’Amico EJ. Density of medical and recreational cannabis outlets: racial/ethnic differences in the associations with young adult intentions to use cannabis, e-cigarettes, and cannabis mixed with tobacco/nicotine. Journal of Cannabis Research. 2021; 3(1): 1 – 9.

7. Stains EL, Kennalley AL, Bachir AS, Kraus CK, Piper BJ. Is medical cannabis evidence-based medicine? Concerns based on qualifying conditions and the National Academy of Sciences Report. MedRxiv [preprint]. doi: https://doi.org/10.1101/2023.05.01.23289286.

8. Boehnke KF, Gangopadhyay S, Clauw DJ, Haffajee RL. Qualifying conditions of medical cannabis license holders in the United States. Health Affairs. 2019; 38(2): 295–302.

9. Cunningham CO, Zhang C, Hollins M, Wang M, Singh-Tan S, Joudrey PJ. Availability of medical cannabis services by racial, social, and geographic characteristics of neighborhoods in New York: a cross-sectional study. BMC Public Health. 2022; 22:671.

10. Cohn AM, Sedani A, Niznik T, Alexander A, Lowery B, McQuoid J, Campbell J. Population and neighborhood correlates of cannabis dispensary locations in Oklahoma. Cannabis. 2023; 6(10): 99–113.

11. Thomas C, Freisthler B. Examining the locations of medical marijuana dispensaries in Los Angeles. Drug Alcohol Rev. 2016; 35(3): 334 – 337.

12. Subica AM, Douglas JA, Kepple NJ, Villanueva S, Grills CT. The geography of crime and violence surrounding tobacco shops, medical marijuana dispensaries, and off-sale alcohol outlets in a large, urban low-income community of color. Preventive Medicine. 2018; 108: 8 – 16.

13. Mair C, Freisthler B, Ponicki WR, Gaidus A. The impacts of marijuana dispensary density and neighborhood ecology on marijuana abuse and dependence. Drug and Alcohol Dependence. 2015; 154: 111 – 116.

14. Freisthler B, Ponicki WR, Gaidus A, Gruenewald PJ. A micro-temporal geospatial analysis of medical marijuana dispensaries and crime in Long Beach California. Addiction. 2016; 111(6): 1027 – 1035.

15. USDA. Rural-Urban Continuum Codes. 2020. https://www.ers.usda.gov/data-products/rural-urban-continuum-codes/. Accessed August 2023.

16. Westreich D, Cole SR. Invited commentary: positivity in practice. American Journal of Epidemiology. 2010; 171(6): 647 – 677.

17. National Conference of State Legislature. State Medical Cannabis Laws. 2023. https://www.ncsl.org/health/state-medical-cannabis-laws#:∼:text=Medical%2DUse%20Update,1%20below%20for%20additional%20information. Accessed August 2023.

18. Boehnke KF, Dean O, Haffajee R, Hosanagar A. US trends in registration for medical cannabis and reasons for use from 2016 to 2020: An observational study. Ann Internal Medicine. 2022; 175(7): 945 – 951.

19. Valencia C, Asaolu IO, Ehiri JE, Rosales C. Structural barriers in access to medical marijuana in the USA – a systematic review protocol. Systematic Reviews. 2017; 65: 154.

20. Mahon E. High Anxiety. Spotlight PA. January 31, 2023. https://www.spotlightpa.org/news/2023/01/pa-medical-marijuana-certification-card-anxiety/. Accessed August 2023.

21. Rosenblatt RA, Andrilla CHA, Catlin M, Larson EH. Geographic and specialty distribution of US physicians trained to treat opioid use disorder. Annals of Family Medicine. 2015; 13(1): 23 – 26.

22. Hollander MAG, Chang CH, Douaihy AB, Hulsey E, Donohue JM. Racial inequity in medication treatment for opioid use disorder: Exploring potential facilitators and barriers to use. Drug and Alcohol Dependence. 2021: 227: https://doi.org/10.1016/j.drugalcdep.2021.108927.

23. Fernandez IS, Stephen C, Loddenkemper T. Disparities in epilepsy surgery in the United States of America. J Neuro. 2017; 264: 1735–1745.

24. Douthit N, Kiv S, Dwolatsky T, Biswas S. Exposing some important barriers to health care access in rural USA. Public Health. 2015; 129: 611 – 620.

25. 2016 Act 16. Chapter 8 Dispensaries. Pennsylvania General Assembly. 2016. https://www.legis.state.pa.us/cfdocs/legis/LI/uconsCheck.cfm?txtType=HTM&yr=2016&sessInd=0&smthLwInd=0&act=016&chpt=8. (Accessed August 2023).

26. Careless R. Zoning hurdles for budding Pennsylvania cannabis businesses. Temple 10-Q. 2023. https://www2.law.temple.edu/10q/zoning-hurdles-budding-pennsylvania-cannabis-businesses/. Accessed August 2023.

27. Von Both I, Santos B. Death of a young woman with cyclic vomiting: a case report. Forensic Science Medical Pathology. 2021; 17(4): 715 – 722.

