## Supplemental Table 1 for "Dispensaries and medical marijuana certifications and indications: Unveiling the geographic connections in Pennsylvania, USA"

**Supplemental Table 1**. Certifying conditions by year in Pennsylvania. Superscript numbers refer to page numbers of the *National Academy of Science* 2017 report “The Health Effects of Cannabis and Cannabinoids.”*

| **2016** | **2017** | **2018** | **2019** | **2020** | **2021** | **2022** | **2023** |
| --- | --- | --- | --- | --- | --- | --- | --- |
| ALS^105^ | ALS^105^ | ALS^105^ | ALS^105^ | ALS^105^ | ALS^105^ | ALS^105^ | ALS^105^ |
|  |  |  | Anxiety disorders^118^ | Anxiety disorders^118^ | Anxiety disorders^118^ | Anxiety disorders^118^ | Anxiety disorders^118^ |
| Autism | Autism | Autism | Autism | Autism | Autism | Autism | Autism |
| Cancer^87,90,91, 94^ | Cancer^87,90,91, 94^ | Cancer^87,90,91, 94^ | Cancer^87,90,91, 94^ | Cancer^87,90,91, 94^ | Cancer^87,90,91, 94^ | Cancer^87,90,91, 94^ | Cancer^87,90,91, 94^ |
| Crohn’s Disease | Crohn’s Disease | Crohn’s Disease | Crohn’s Disease | Crohn’s Disease | Crohn’s Disease | Crohn’s Disease | Crohn’s Disease |
| Damage to the nervous tissue of the spinal cord with objective neurological indication of intractable spasticity^101^ | Damage to the nervous tissue of the spinal cord with objective neurological indication of intractable spasticity^101^ | Damage to the nervous tissue of the spinal cord with objective neurological indication of intractable spasticity^101^ | Damage to the nervous tissue of the spinal cord with objective neurological indication of intractable spasticity^101^ | Damage to the nervous tissue of the spinal cord with objective neurological indication of intractable spasticity^101^ | Damage to the nervous tissue of the spinal cord with objective neurological indication of intractable spasticity^101^ | Damage to the nervous tissue of the spinal cord with objective neurological indication of intractable spasticity^101^ | Damage to the nervous tissue of the spinal cord with objective neurological indication of intractable spasticity^101^ |
|  |  | Dyskinetic and spastic movement disorders^101,110^ | Dyskinetic and spastic movement disorders^101,110^ | Dyskinetic and spastic movement disorders^101,110^ | Dyskinetic and spastic movement disorders^101,110^ | Dyskinetic and spastic movement disorders^101,110^ | Dyskinetic and spastic movement disorders^101,110^ |
| Epilepsy^99^ | Epilepsy^99^ | Epilepsy^99^ | Epilepsy^99^ | Epilepsy^99^ | Epilepsy^99^ | Epilepsy^99^ | Epilepsy^99^ |
| Glaucoma^113^ | Glaucoma^113^ | Glaucoma^113^ | Glaucoma^113^ | Glaucoma^113^ | Glaucoma^113^ | Glaucoma^113^ | Glaucoma^113^ |
| HIV/AIDS^94^ | HIV/AIDS^94^ | HIV/AIDS^94^ | HIV/AIDS^94^ | HIV/AIDS^94^ | HIV/AIDS^94^ | HIV/AIDS^94^ | HIV/AIDS^94^ |
| Huntington’s Disease^106^ | Huntington’s Disease^106^ | Huntington’s Disease^106^ | Huntington’s Disease^106^ | Huntington’s Disease^106^ | Huntington’s Disease^106^ | Huntington’s Disease^106^ | Huntington’s Disease^106^ |
| Inflammatory Bowel Disease | Inflammatory Bowel Disease | Inflammatory Bowel Disease | Inflammatory Bowel Disease | Inflammatory Bowel Disease | Inflammatory Bowel Disease | Inflammatory Bowel Disease | Inflammatory Bowel Disease |
| Intractable seizures^99^ | Intractable seizures^99^ | Intractable seizures^99^ | Intractable seizures^99^ | Intractable seizures^99^ | Intractable seizures^99^ | Intractable seizures^99^ | Intractable seizures^99^ |
| Multiple Sclerosis^101^ | Multiple Sclerosis^101^ | Multiple Sclerosis^101^ | Multiple Sclerosis^101^ | Multiple Sclerosis^101^ | Multiple Sclerosis^101^ | Multiple Sclerosis^101^ | Multiple Sclerosis^101^ |
|  |  | Neurodegenerative diseases | Neurodegenerative diseases | Neurodegenerative diseases | Neurodegenerative diseases | Neurodegenerative diseases | Neurodegenerative diseases |
| Neuropathies^87^ | Neuropathies^87^ | Neuropathies^87^ | Neuropathies^87^ | Neuropathies^87^ | Neuropathies^87^ | Neuropathies^87^ | Neuropathies^87^ |
|  |  | Opioid Use Disorder (if conventional therapeutic interventions are contraindicated or ineffective, or adjunctive therapy)^116^ | Opioid Use Disorder (if conventional therapeutic interventions are contraindicated or ineffective, or adjunctive therapy)^116^ | Opioid Use Disorder (if conventional therapeutic interventions are contraindicated or ineffective, or adjunctive therapy)^116^ | Opioid Use Disorder (if conventional therapeutic interventions are contraindicated or ineffective, or adjunctive therapy)^116^ | Opioid Use Disorder (if conventional therapeutic interventions are contraindicated or ineffective, or adjunctive therapy)^116^ | Opioid Use Disorder (if conventional therapeutic interventions are contraindicated or ineffective, or adjunctive therapy)^116^ |
| Parkinson’s Disease^108^ | Parkinson’s Disease^108^ | Parkinson’s Disease^108^ | Parkinson’s Disease^108^ | Parkinson’s Disease^108^ | Parkinson’s Disease^108^ | Parkinson’s Disease^108^ | Parkinson’s Disease^108^ |
| PTSD^123^ | PTSD^123^ | PTSD^123^ | PTSD^123^ | PTSD^123^ | PTSD^123^ | PTSD^123^ | PTSD^123^ |
| Sickle Cell Anemia | Sickle Cell Anemia | Sickle Cell Anemia | Sickle Cell Anemia | Sickle Cell Anemia | Sickle Cell Anemia | Sickle Cell Anemia | Sickle Cell Anemia |
| Severe chronic or intractable pain^87^ | Severe chronic or intractable pain^87^ | Severe chronic or intractable pain^87^ | Severe chronic or intractable pain^87^ | Severe chronic or intractable pain^87^ | Severe chronic or intractable pain^87^ | Severe chronic or intractable pain^87^ | Severe chronic or intractable pain^87^ |
|  |  | Terminal illness | Terminal illness | Terminal illness | Terminal illness | Terminal illness | Terminal illness |
|  |  |  | Tourette syndrome^103^ | Tourette syndrome^103^ | Tourette syndrome^103^ | Tourette syndrome^103^ | Tourette syndrome^103^ |

| KEY | |
| --- | --- |
| Green | There is conclusive or substantial evidence that cannabis or cannabinoids are effective for this condition |
| Yellow | There is moderate or limited evidence that cannabis or cannabinoids are effective for this condition |
| Red | There is limited evidence that cannabis or cannabinoids are *ineffective* for this condition, or there is no or insufficient evidence to support or refute the conclusion that cannabis or cannabinoids are an effective treatment for this condition |
| Blue | This condition or its symptoms are mentioned in the NAS Report, but that which the state lists as a condition doesn’t fit well into a studied category |
| Gray | This condition is not mentioned in the NAS Report |
